# Previous experience with depression in others and self: a cross-sectional study of the impact on stigma and help-seeking attitudes

**DOI:** 10.1101/2020.05.02.20086140

**Authors:** Virgínia da Conceição, Inês Rothes, Milton Severo, Kathy Griffiths, Ulrich Hegerl, Ricardo Gusmão

**Affiliations:** EPIUnit - Institute of Public Health, University of Porto, Portugal; Faculty of Psychology and Education Science, University of Porto, Portugal; Center for Psychology at University of Porto, Portugal; Department of Public Health and Forensic Sciences, and Medical Education, Faculty of Medicine, University of Porto, Portugal; Research School of Psychology, College of Health & Medicine, The Australian National University, Australia; Department of Psychiatry, Psychosomatics, and Psychotherapy, Goethe-Universität Frankfurt, Frankfurt, Germany

**Keywords:** Depression, stigma, help-seeking behaviour

## Abstract

**Background:** Stigma has been considered a significant barrier both in treatment, rehabilitation and help-seeking behaviours of people diagnosed with depression. This study aimed to assess the influence of the type of previous experience with depression on depression stigma, identify the effects of previous experience with depression on stigma and to analyse the effects of stigma on help-seeking attitudes.

**Methods:** A total of 1693 participants with a mean age of 47.2 (SD=18.17) completed the Depression Stigma Scale (DSS), the Attitude Toward Seeking Professional Psychological Help (ATSPPH), and a sociodemographic questionnaire. We categorised participants into four comparison groups: no previous experience with depression (n=479), indirect experience with depression (n=661), direct experience with depression (n=137), and both direct and indirect experience with depression (n=416). Data were analysed using SPSS 24.0.

**Results:** Levels of personal stigma were lower in people who had family and friends experiencing depression in comparison with individuals with no history of depression experience. Better attitudes towards help-seeking were evident in those with lower personal stigma, and worse help-seeking attitudes were associated with higher perceived stigma in the indirect previous experience group.

**Limitations:** Duration of participant exposure to depression was not collected.

**Conclusions:** The individual’s experience with depression influences the development of personal stigmatisation towards depression and plays a role in help-seeking behaviours. Addressing people’s experience of depression might be a practical way of reducing depression stigma and improve help-seeking behaviours.

## 1. Introduction

Globally, people diagnosed with depressive disorders comprise 4.4% of world population, and depression ranks as the most significant contributor to non-fatal health loss, accounting for 7.5% of all Years Lived with Disability (YLD) (1). Moreover, although the underlying mechanisms are unclear, there is a well-documented association between depression and lower life expectancy (2) corroborated in a recent longitudinal population-based study which reported a higher rate of mortality over an 11-year follow-up period for individuals with psychological distress or major depressive disorder (3).

Along with this high level of global burden, the treatment gap for people with depressive disorders worldwide is very significant, indicating that the majority of people do not access or receive the medical and psychological care they need (4) despite the well-established return on investment associated with treatment (5).

One barrier to care access and adequate treatment might be the stigma associated with mental illness. There is evidence that such stigma, which is associated with feelings of shame, guilt, lack of self-esteem and social isolation (6, 7) as well as physical effects such as hypertension, elevated cortisol levels, and experiences of chronic pain (8), negatively influences help-seeking and the therapeutic process (6, 8-14). Thus, reducing stigma affecting people who have a mental illness such as depression is one of the main objectives of the Mental Health Action Plan 2013-2020 of the World Health Organization (15).

Stigma is a major issue for people with a mental illness, including those living with depression. For example, people with depression have reported community and institutional stigma similar to that experienced by people with psychosis (16) and a recent study found that 79% of the participants with major depressive disorders had experienced discrimination associated with their condition and between 20% and 37% of the participants have compromised their actions because of anticipated discrimination (7).

Much of the recent research on depression and stigma has focused on two types of stigma: personal stigma and perceived stigma, personal stigma referring to one’s own beliefs about depression, and perceived stigma alluding to one’s beliefs about the attitudes of others (9). Recent studies have identified previous experience with depression as one of the leading predictors of personal stigma: a lower level of previous experience is associated with a higher level of personal stigma. In particular, studies have shown that participants with no previous experience with depression obtained greater personal stigma scores than those with previous experience with depression (17). People with a history of depression showed less personal stigma than those with antecedents of parental depression and no previous experience with depression (9). In general, personal stigma decreases with depressive experience magnitude, with no significant impact on perceived stigma (11).

Other identified predictors of personal depression stigma are lower education (18), and gender, with higher scores among male than females (9, 17).

The association between prior experience with depression and perceived stigma is less clear, but some studies have reported some association between prior experience with depression and higher perceived stigma (11).

Stigma plays an essential role in help-seeking behaviours. As it represents an essential vehicle for exploring and understanding patient delay and early action across a variety of health conditions including mental health care access (10), understanding stigma effects is crucial. Even though the importance of understanding the factors that influence help-seeking behaviours is well recognised, there remains a lack of research on the topic (14).

A better understanding of stigma components and its predictors has the potential of significantly improve stigma reduction interventions and promote help-seeking behaviours to decrease unmet need for treatment (9, 11, 19-21).

The aims of this study were therefore threefold: to assess the influence of the type of previous experience with depression on the levels of depression stigma in different sociodemographic groups, and to identify the relationship between depression stigma and help-seeking behaviour according to the type of previous experience with depression and its value as a predictor for help-seeking.

## 2. Methods

### 2.1 Participants and procedures

The sample collection for this study was part of the OSPI program – Optimising suicide prevention programs and their implementation in Europe, specifically within the application in Portugal (12, 22), and the current study assumed a cross-sectional study design.

In Portugal, selection of participants was from the cable telephone network listed numbers, using the random digit dialling method to numbers belonging to Almada and Amadora municipalities. Trained interviewers conducted telephone contacts.

Almada and Amadora combined have above 350 000 inhabitants, being two of the most populous counties in Portugal. Due to its high population density, and the socioeconomic diversity of its inhabitants, these municipalities were considered representative of the Portuguese population (22).

Of the 2009 participants in the OSPI research program (22, 23), we excluded 316 participants from the current study due to their exposure to the OSPI intervention which aimed to promote literacy on depression and, consequently, help-seeking attitudes. We collected the data between 2009 and 2010.

Previous experience with depression was self-reported by the participants upon two direct questions (described in the supplementary file).

To investigate the relationship between past contact and stigma and help-seeking, we categorised the remaining 1693 participants into four groups according to their level of past experience of depression. Group A (No experience) included 479 participants without experience of depression in themselves or close contact. Group B (Indirect experience) comprised 661 participants who reported previous indirect experience of depression in a relative or close friend but not themselves. We allocated the 137 participants who had experienced depression themselves to group C (Direct experience). Group D (Direct and Indirect experience) included the 416 participants who had both experienced depression and had a relative or close friend with depression. More information can be found in the supplementary file.

### 2.2 Instruments

Participants completed a short sociodemographic questionnaire, the Portuguese translation of the Depression Stigma Scale (DSS) (24), and the Attitude Toward Seeking Professional Psychological Help: short form (ATSPH) (25). The DSS comprises a 9-item Personal subscale and 9-item Perceived subscale with each item scored on a 5-point Likert scale ranging from 0 (strongly disagree) to 4 (strongly agree); higher scores indicate greater stigma. The ATSPH is a 10-item (range 0 to 30) with higher scores reflecting better help-seeking attitudes.

The sociodemographic questionnaire contained questions concerning the respondent’s sex, age, professional occupation and a question about previous depression diagnosis, a multiple-choice question with options to record the participant’s own experience and that of close relatives and friends (see supplementary file). As described above, we coded the last item into the four categories: no experience of depression, indirect experience, direct experience, and both direct and indirect experience.

### 2.3 Data analysis

In addition to computing a continuous score for each DSS and ATSPPH item, we transformed the DSS score into percentages, in accordance to the original scale (26), with higher percentages indicating greater stigma levels. In the ATSPPH, the higher scores meaning positive help-seeking attitudes.

We also calculated means and standard deviations for continuous variables and undertook ANOVAs followed by the Tukey HSD’s multiple comparisons test to compare stigma and help-seeking levels across groups. We applied Student t-tests to examine the differences between sexes within each contact group, and Pearson correlations to examine the relationship between DSS scores and age.

We performed hierarchical linear regression analyses to identify which variables were significant predictors of the depression stigma and help-seeking behaviour attitudes and the magnitude of the estimations. In the analysis of the predictors of depression stigma, gender and age were entered in the first model (Model 1), occupation in the second (Model 2), and depression experience in the final step (Model 3). In the analysis in which help-seeking was the criterion variable, depression experience was entered in the first step (Model 1), personal stigma in the second (Model 2) and perceived stigma in the final step (Model 4).

Sensitivity analysis was applied to the sample through stratified randomisation according to occupation and analysis repeated.

IBM SPSS Statistics, Version 24.0 software package was used to conduct statistical analysis.

### 2.3 Ethical considerations

Trained interviewers explained in the first contact the objectives of the study and obtained verbal informed consent. Anonymity and confidentiality of the data collected were guaranteed.

The approval of the study protocol occurred in the scope of the OSPI program (12, 23). In Portugal, the OSPI study was approved by the Medical Sciences Faculty of the New University of Lisbon’s ethics commission in May 2009.

## 3. Results

### 3.1 Sample description

As to gender and age, 53.6% were women, and the mean age was 47.2 years (SD=18.17, range 18-90).

In Group A, participants had a mean age of 47.59 (SD=19.81), 42.2% were female; in Group B participants had a mean age of 44.31 (SD=17.52), and 51.7% were female; Group C presented a mean age of 55.49 (SD=17.56) and 64.9% female; finally, participants in Group D had a mean age of 49.15 (SD=16.29), and 66.8% were female (see the table in the supplementary file for more participant characteristics).

As the data used in this study was previously treated and used in other researches – as mentioned above – no missing data was detected in the dataset used.

### 3.2 Personal Depression Stigma

As shown in Table 1, we observed significant differences in personal stigma scores between groups A and B (p<0.001), groups A and D (p<0.001), B and C (p<0.05), and C and D (p<0.001), with the lower means obtained in group D, followed by group B. We found a similar pattern of between-group effects for the female sub-sample.

**Table 1:** Personal depression stigma as a function of previous experience and sex.

|  | Group A<br>(No) | Group B<br>(Indirect) | Group C<br>(Direct) | Group D<br>(Both) | ANOVA<sub>(df)</sub> |
| --- | --- | --- | --- | --- | --- |
| Mean (SD) | 56.72 (11.51) | 38.04 (13.29) | 41.95 (14.40) | 36.14 (14.40) | **F<sub>(3,1689)</sub> = 27.78**<br>**p<0.001** |
| Male<br>Mean (SD) | 44.88 (13.63) | 38.99 (12.96) | 39.73 (14.88) | 38.57 (15.07) | **F<sub>(3,782)</sub> = 11.26,**<br>**p<0.001** |
| Female<br>Mean (SD) | 42.72 (13.21) | 37.14 (13.54) | 43.14 (14.08) | 34.94 (13.92) | **F<sub>(3,903)</sub> = 16.83**<br>**p<0.001** |
| t-test<sub>(df)</sub> | t<sub>(472)</sub>=1.73,<br>p=0.08 | t<sub>(689)</sub>= 1.83,<br>p=0.07 | t<sub>(129)</sub>= -1.29,<br>p=0.19 | **t<sub>(395)</sub>= 2.39,**<br>**p<0.05** |  |
| Previous experience*sex |  |  |  |  | F<sub>(3,1689)</sub> = 1.99<br>p=0.13 |

M=mean, SD=standard deviation; df=degrees of freedom. Significant results are shown in bold.

Gender differences were significant in group D, where males showed higher personal stigma means than females. There was a significant positive correlation between age and personal stigma in each of the previous experience of depression groups (group A: r=0.37, p <0.001; group B: r=0.30, p <0.001; group C: r=0.22, p <0.05; and group D: r=0.22, p <0.001), the higher the age, the higher the personal stigma. Personal stigma was higher among retired participants than any other occupation (M=46.43, SD=14.35; F=54.12, p<0.001). Pearson correlations are statistically different between group A and group C (z=1.69, p<0.05) and between group A and group D (z=2.45, p<0.01).

Hierarchical linear regression analysis (Table 2) indicated that gender and age were significant predictors of depression personal stigma after controlling for all other factors. Being employed has a significant effect on personal depression stigma reduction. Controlling for other factors, participants with previous direct and indirect experience of depression (Group D) showed significantly lower personal stigma than participants from other groups, with the strongest effect for those with no experience of depression (Group A)

**Table 2:**
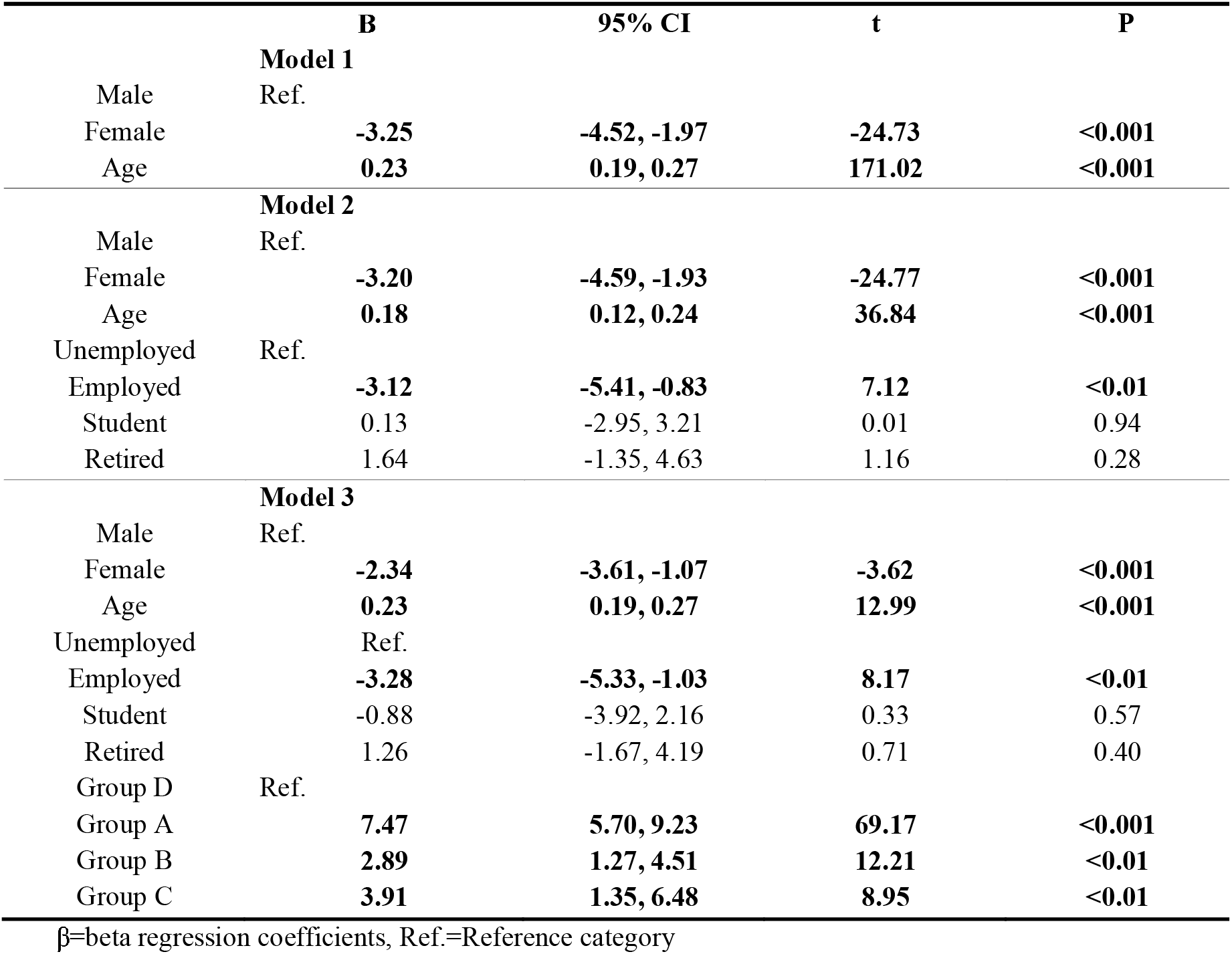
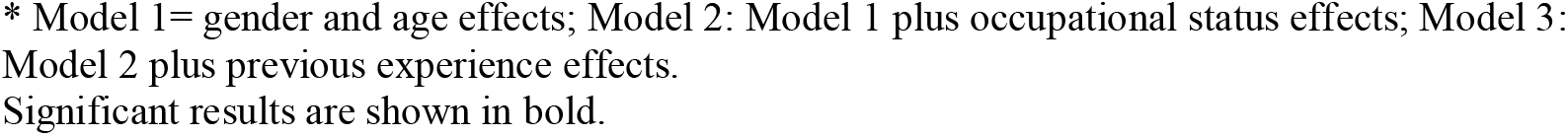
Regression models on the different types of experience, gender, age and occupation among personal depression stigma

|  | B | 95% CI | t | P |
| --- | --- | --- | --- | --- |
| **Model 1** |  |  |  |  |
| Male | Ref. |  |  |  |
| Female | **-3.25** | **-4.52, -1.97** | **-24.73** | **<0.001** |
| Age | **0.23** | **0.19, 0.27** | **171.02** | **<0.001** |
| **Model 2** |  |  |  |  |
| Male | Ref. |  |  |  |
| Female | **-3.20** | **-4.59, -1.93** | **-24.77** | **<0.001** |
| Age | **0.18** | **0.12, 0.24** | **36.84** | **<0.001** |
| Unemployed | Ref. |  |  |  |
| Employed | **-3.12** | **-5.41, -0.83** | **7.12** | **<0.01** |
| Student | 0.13 | -2.95, 3.21 | 0.01 | 0.94 |
| Retired | 1.64 | -1.35, 4.63 | 1.16 | 0.28 |
| **Model 3** |  |  |  |  |
| Male | Ref. |  |  |  |
| Female | **-2.34** | **-3.61, -1.07** | **-3.62** | **<0.001** |
| Age | **0.23** | **0.19, 0.27** | **12.99** | **<0.001** |
| Unemployed | Ref. |  |  |  |
| Employed | **-3.28** | **-5.33, -1.03** | **8.17** | **<0.01** |
| Student | -0.88 | -3.92, 2.16 | 0.33 | 0.57 |
| Retired | 1.26 | -1.67, 4.19 | 0.71 | 0.40 |
| Group D | Ref. |  |  |  |
| Group A | **7.47** | **5.70, 9.23** | **69.17** | **<0.001** |
| Group B | **2.89** | **1.27, 4.51** | **12.21** | **<0.01** |
| Group C | **3.91** | **1.35, 6.48** | **8.95** | **<0.01** |

β=beta regression coefficients, Ref.=Reference category
\* Model 1= gender and age effects; Model 2: Model 1 plus occupational status effects; Model 3: Model 2 plus previous experience effects. Significant results are shown in bold.

### 3.4 Perceived Depression Stigma

There were no significant differences in perceived stigma as a function of the type of experience of depression or gender, except perceived stigma was lower among male than female participants in Group D (Table 3).

**Table 3:**
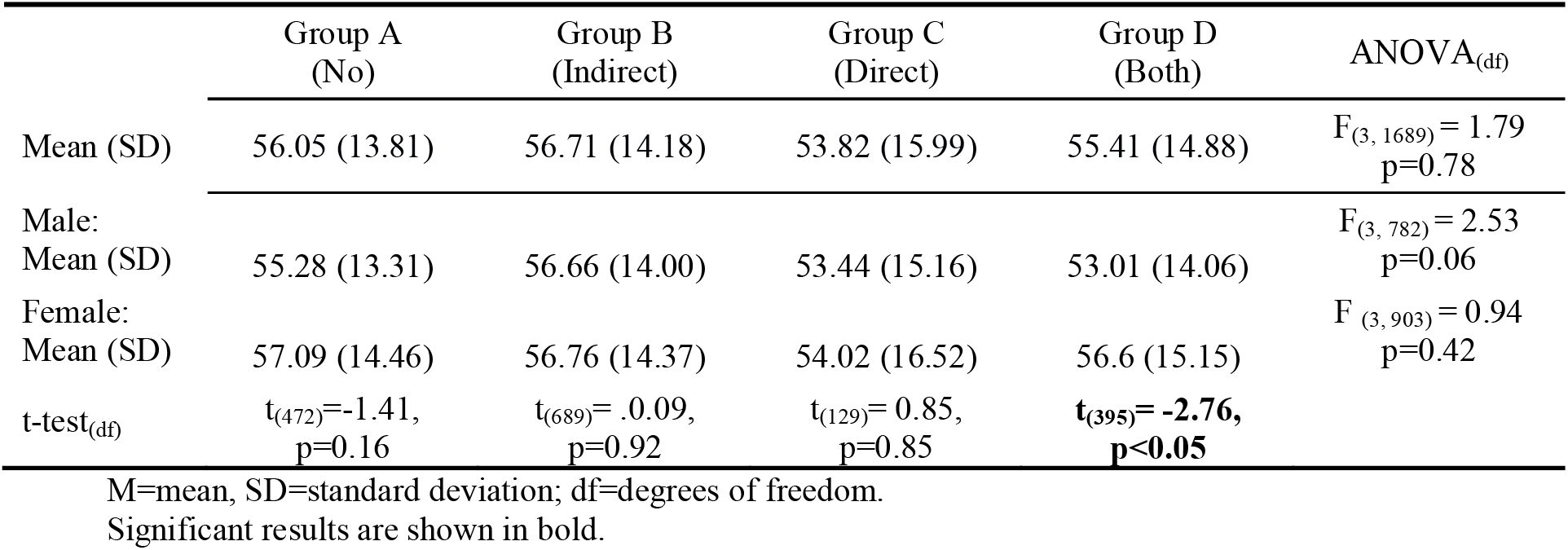
Perceived depression stigma as a function of different types of previous experience and sex.

There was a statistically significant, albeit weak, negative correlation between age and perceived stigma for each of the previous experience of depression groups: group A r=-0.107, p <0.05; group B r=-0.130, p <0.05; group C r=-0.176, p <0.05; and group D r=-0.192, p <0.001.

There was also a main effect of occupational group on perceived stigma (F=12.96, p<0.001), with all pairwise groups comparisons reaching statistical significance except the contrast between employed and unemployed participants. Students had the highest level of perceived stigma (M=61.23, SD=12.03) and retired participants the lowest (M=53.41, SD=14.81).

A hierarchical regression analysis confirmed that previous experience with depression was not a significant independent predictor of perceived depression stigma (see Supplementary Table). After controlling for other variables, only gender and age were predictors of perceived stigma with perceived stigma being higher among females than males (β=1.42, t=4.16, p<0.05) and older age associated with lower perceived stigma (β=-0.12, t=38.87, p<0.001).

### 3.5 Help-seeking attitudes

Personal stigma has a significant impact on help-seeking attitudes, with an r=-0.339, p <0.001; however, we found no significance on the correlation between perceived stigma and help-seeking attitudes.

As shown in Table 4, analysis of variance followed by paired comparisons indicated there was a statistically significant difference in the help-seeking attitudes between group A and group B with the group with previous indirect experience having more favourable attitudes to help-seeking than the group without the experience of depression. The females in groups A and D showed more favourable attitudes to help-seeking than their male counterparts, as shown in Table 4. Less positive help-seeking attitudes were weakly associated with older age in each group except group C (group A: r=-0.190, p <0.001; group B r=-0.08, p <0.05; group C r=-0.04, p = 0.65; and group D r=-0.105, p <0.05).

**Table 4:** Help-seeking attitudes as a function of previous experience and sex.

|  | Group A<br>(No) | Group B<br>(Indirect) | Group C<br>(Direct) | Group D<br>(Both) | ANOVA<sub>(df)</sub> |
| --- | --- | --- | --- | --- | --- |
| Mean (SD) | 17.49 (5.0) | 18.57 (4.99) | 17.80 (5.14) | 18.20 (5.13) | **$F_{(3, 1689)} = 4.59$**<br>**$p<0.01$** |
| Male:<br>Mean (SD) | 16.85 (5.08) | 18.26 (5.13) | 17.33 (6.42) | 17.10 (5.55) | **$F_{(3, 782)} = 4.01$**<br>**$p<0.01$** |
| Female:<br>Mean (SD) | 18.36 (4.78) | 18.87 (4.86) | 18.06 (4.32) | 18.75 (4.83) | $F_{(3, 903)} = 0.96$<br>$p=0.41$ |
| t-test<sub>(df)</sub> | **$t_{(472)}=-3.28,$**<br>**$p<0.01$** | $t_{(689)}= -1.58,$<br>$p=0.11$ | $t_{(129)}= -0.69,$<br>$p=0.44$ | **$t_{(395)}= -292,$**<br>**$p<0.01$** | |
| Previous experience*sex | | | | | $F_{(3, 1689)} = 1.21$<br>$p=0.30$ |

M=mean, SD=standard deviation; df=degrees of freedom. Significant results are shown in bold.

Considering the model that included only the type of previous experience with depression in the hierarchical regression analysis (Table 5), groups B and D demonstrated better attitudes towards help-seeking than their counterparts with no previous experience of depression. However, after controlling for personal stigma (Model 2), the effect of previous experience was no longer significant, and higher levels of personal stigma predicted less positive help-seeking attitudes (see Table 5). Perceived stigma did not add significantly to the model (Model 4).

**Table 5:**
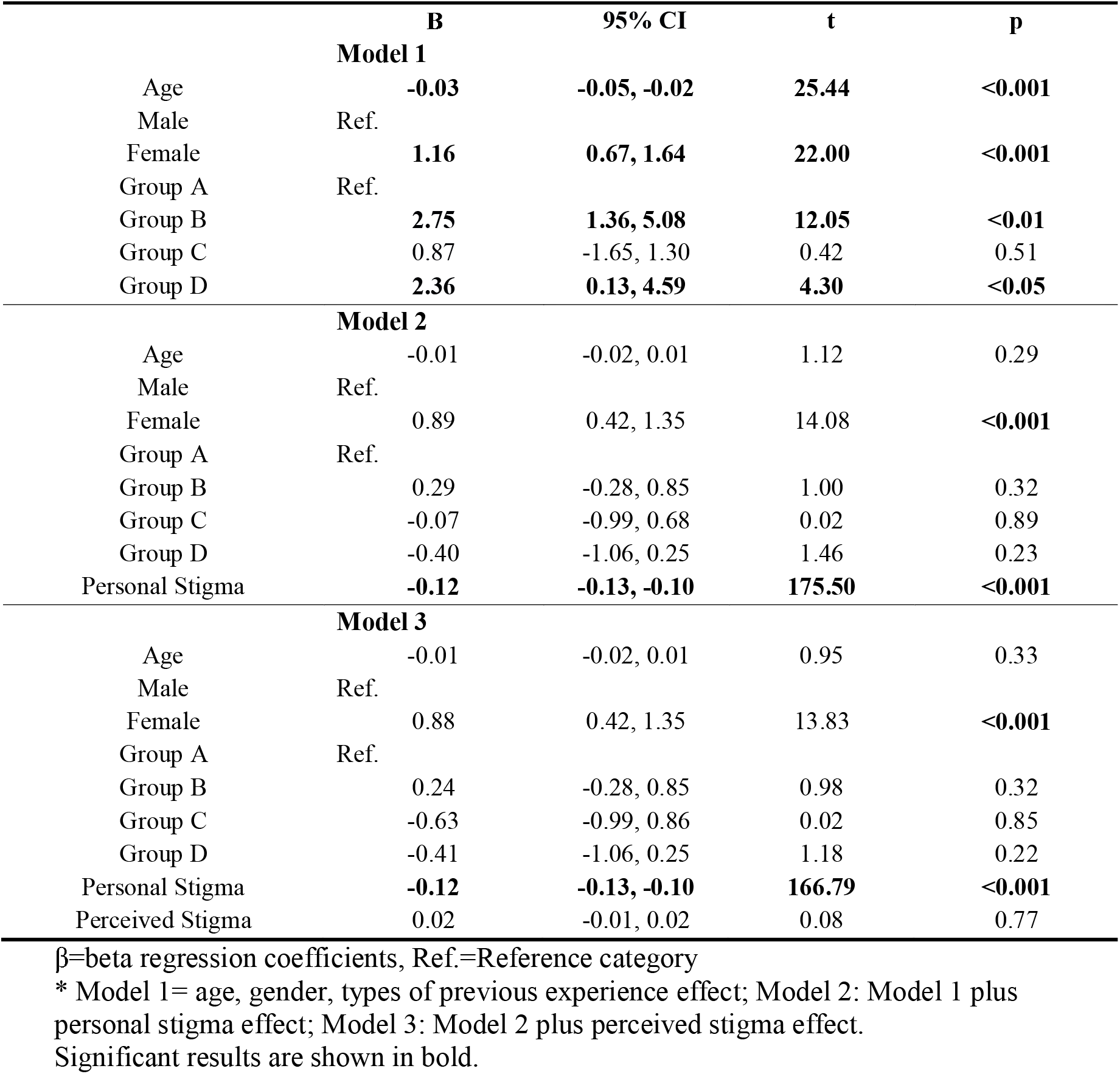
Effects of age, gender, different types of previous experience, personal and perceived stigma on help-seeking behaviour.

As shown in Table 6, personal stigma had a negative effect on help-seeking in all groups, proving to be a strong predictor independently of the type of previous experience with depression. On the other hand, perceived stigma was a significant predictor only for those with indirect experience of depression (group B) with the higher perceived stigma associated with less positive attitudes to help-seeking in this group.

**Table 6:**
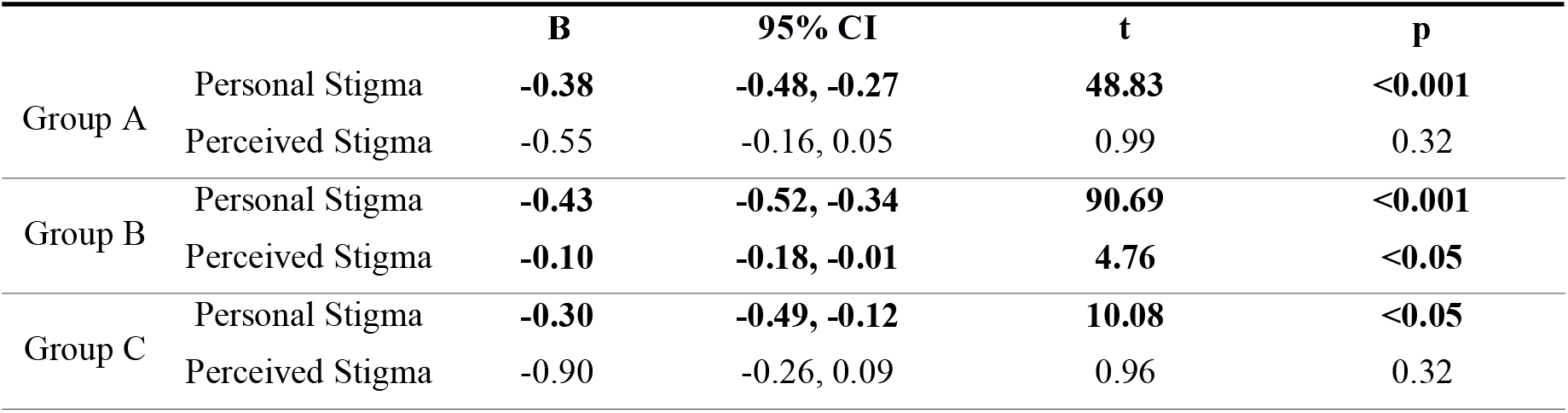
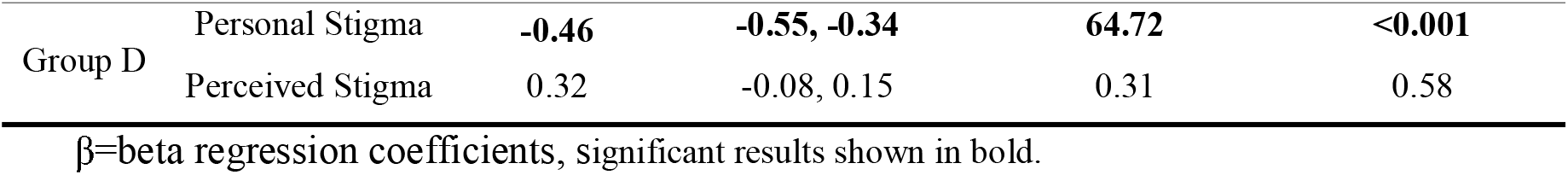
Type of previous experience effects on the relationship between stigma and help-seeking attitudes.

|  |  | **B** | **95% CI** | **t** | **p** |
| --- | --- | --- | --- | --- | --- |
| Group A | Personal Stigma | **-0.38** | **-0.48, -0.27** | **48.83** | **<0.001** |
|  | Perceived Stigma | -0.55 | -0.16, 0.05 | 0.99 | 0.32 |
| Group B | Personal Stigma | **-0.43** | **-0.52, -0.34** | **90.69** | **<0.001** |
|  | Perceived Stigma | **-0.10** | **-0.18, -0.01** | **4.76** | **<0.05** |
| Group C | Personal Stigma | **-0.30** | **-0.49, -0.12** | **10.08** | **<0.05** |
|  | Perceived Stigma | -0.90 | -0.26, 0.09 | 0.96 | 0.32 |
| Group D | Personal Stigma | **-0.46** | **-0.55, -0.34** | **64.72** | **<0.001** |
|  | Perceived Stigma | 0.32 | -0.08, 0.15 | 0.31 | 0.58 |

$\beta$ =beta regression coefficients, significant results shown in bold.

### 3.6 Sensitivity analysis

Due to the occupational differences between Group C and other groups, we collected a more similar subsample in the other groups using the stratified randomisation method with the occupational status as the stratification factor. This procedure resulted in a sample of 1053 individuals: 316 in group A, with a mean age of 54.1 (SD=19.07) and 42.1% females; 318 in group B, 54.0 (SD=18.95) years of age where 55.7% were female; and 288 in group D, 52.4 (17.48) years old and 66.7% female. A more detailed description of the subsample can be found in the supplementary file.

Personal depression stigma in this subsample in descending order was as follows: group A (M=45.54, SD=14.08), group C (M=41.95, SD=14.40), group B (M=40.26, SD=13.58), and group D (M=36.80, SD=14.74). There was a significant difference in personal stigma levels across groups (F(3,1049) = 19.72 p<0.001). The level of personal stigma in group D (direct and indirect experience) was lower than in each of the other groups (group A: p<.001; groups B, C: p<.05) and personal stigma in group B (indirect experience) was lower than in group A (no experience) (p<0.05).

In this subsample, there were no significant differences in perceived stigma as a function of the type of previous experience (F(3,1049) = 1.28, p=0.28) with mean perceived stigma ranging from 53.82 (SD=16.00) in group C to 56.71 (SD=14.18) in group B. As in the total sample, the only difference observed was between genders in group D (t_(287)_= −2.66, p<0.05).

## 4. Discussion

### 4.1 Summary of main findings

The first aim was to examine the influence of previous experience with depression on personal and perceived depression stigma. The absence of previous experience of depression (Group A) was associated with the highest level of personal stigma. Interestingly, having experienced depression oneself (Group C) was associated with higher personal depression stigma than with watching friends and family members suffer the disorder (Group B). People who had experienced depression both directly and indirectly (group D) showed lower levels of personal stigma than any other group, followed by those with close contact with a person with depression (Group B). In contrast, the type of previous experience with depression was not a predictor of perceived stigma, except for gender differences.

The second aim of the study was to examine the relationship between depression stigma and help-seeking behaviour according to the type of previous experience with depression and the value of personal and perceived stigma as a predictor of help-seeking. We found a negative effect of personal depression stigma on help-seeking attitudes for every type of previous experience with depression, whereas for perceived depression stigma effects were non-significant, except for females.

### 4.2 Discussion and literature

Although several studies have explored the effects of previous experience with mental disorders on stigma (11), and on depression in particular (9, 27), to our knowledge, the current study is the first to report systematically on the gradient of exposure to depression.

#### 4.2.1 Personal Depression stigma

The finding of lower levels of personal stigma among those experiencing depression in other or self is consistent with previous studies that have reported a negative association between level of previous experience with depression and personal stigma (9, 11). However, the previous indirect experience group (B) had a significantly lower personal stigma mean than direct previous experience (C).

To our knowledge, no other research has compared the personal stigma between the direct and indirect previous experience with depression.

The gender differences observed in people in Group D who had experienced depression both directly and indirectly are in line with the findings of previous studies, but there were no significant observable differences in the other groups. In other words, in groups with a higher personal stigma, no gender differences were detected, suggesting that the experience of depression has a more substantial effect on females.

Within the male group, individuals with no previous experience with depression (A) had higher personal stigma than those with previous indirect experience (B) and both previous direct and indirect experience (D). However, previous direct experience (C) had no significant effect on personal stigma reduction. We found similar results within the female group; yet, in this case, females with direct experience (C) showed the second-highest personal stigma means.

These results may provide an essential insight into the effect of type of depression experience on stigma. It is possible that observing significant pain in someone close reduces personal depression stigma more than the more proximal experience of suffering depression oneself with individuals being more sympathetic and less judgmental about others than with themselves.

On the other hand, ageing has a stronger effect on stigmatisation when people have experienced depression through a close other (group B) than when experiencing depression oneself (group C), raising the hypotheses of evanescent recall. Future research should take into account the time elapsed since the depression experience because it would be interesting to understand the depression stigma reduction durability of the previous experience with depression.

It is legitimate to speculate that some internalisation of the discrimination experiences reported in previous studies (7) may have occurred and therefore, working on stigma reduction among people with depression may be of particular clinical relevance.

#### 4.2.2 Perceived Depression stigma

On the other hand, the type of previous experience of depression was not a necessary predictor of perceived stigma. However, men experiencing depression in self and others (group D) presented less perceived stigma, then those who did not experience it (A) or experienced it only in others (B). There were also gender differences in both types of previous experience group (group D), with a higher mean perceived stigma among females. The explanation for these findings is unclear but suggests that when experiencing depression in self and others, men feel others are less stigmatising, externalising their feelings.

#### 4.2.3 Personal versus perceived stigma

In this study, we found differences between personal and perceived stigma. Despite a slightly positive correlation between these two subscales, whereas personal stigma increased with age, perceived stigma diminished with age. Also, women presenting a lower score in personal stigma show a higher score in perceived stigma.

We observed a positive correlation between age and personal depression stigma, with higher scores in older individuals, in agreement with previous results (Boerema et al., 2016) and the opposite effect on perceived stigma. This correlation was highest in individuals with no previous experience of depression for personal stigma when comparing with those with direct and both direct and indirect previous experience with depression.

Thus, there is a suggestion of an inverse correlation between these two constructs: the lower the personal stigma, the higher the perceived stigma, as previously reported in the literature (9). It is also possible that perceived stigma scores reflect an oversensitivity and subsequently a resulting overestimation of the stigma of the community by those with lower levels of personal stigma.

#### 4.2.4 Help-seeking

Personal depression stigma had a significant adverse effect on help-seeking attitudes in every group, in agreement with previous studies (28). On the other hand, perceived stigma bear no effect on help-seeking attitudes in our study, contrary to what was previously observed in previous studies (29). Except for those with previous indirect experience, where we observed a decrease in the likelihood of seeking professional help from those with higher perceived stigma.

#### 4.2.7 Limitations

There are several limitations to this study.

Firstly, there are some differences in the sociodemographic characteristics across groups, namely group C. Secondly, phone interviews may have induced some socially desired answers. Thirdly, there is no information on the severity, duration and time of onset of the depressive episode, which could be new study variables for further research. Fourthly, the cross-sectional nature of the study was a constraining factor limiting the precluding the exploration of other variables that could affect stigma levels, such as mental literacy promotion over time, the use of mental health care services, and experiences of discriminatory attitudes. Finally, uncontrolled depression severity at the time of assessment could influence the perceived stigma. Future studies may benefit from its inclusion both to better understand the effects as well as to control it as a possible cofounder. These limitations deserve future consideration in further research.

## Data Availability

Data is available upon request to the corresponding author.

